# Evaluation of a Commercial Culture-free Neutralization Antibody Detection Kit for Severe Acute Respiratory Syndrome-Related Coronavirus-2 and Comparison with an Anti-RBD ELISA Assay

**DOI:** 10.1101/2021.01.23.21250325

**Authors:** Jesse Papenburg, Matthew P. Cheng, Rachel Corsini, Chelsea Caya, Emelissa Mendoza, Kathy Manguiat, L. Robbin Lindsay, Heidi Wood, Michael A. Drebot, Antonia Dibernardo, Gerasimos Zaharatos, Reneée Bazin, Romain Gasser, Mehdi Benlarbi, Gabrielle Gendron-Lepage, Guillaume Beaudoin-Bussières, Jérémie Prévost, Andrés Finzi, Momar Ndao, Cedric P Yansouni

## Abstract

**Background:** SARS-CoV-2 surrogate neutralization assays that obviate the need for viral culture offer substantial advantages regarding throughput and cost. The cPass SARS-CoV-2 Neutralization Antibody Detection Kit (Genscript) is the first such commercially available assay, detecting antibodies that block RBD/ACE-2 interaction. We aimed to evaluate cPass to inform its use and assess its added value compared to anti-RBD ELISA assays.

**Methods:** Serum reference panels comprising 205 specimens were used to compare cPass to plaque-reduction neutralization test (PRNT) and a pseudotyped lentiviral neutralization (PLV) assay for detection of neutralizing antibodies. We assessed the correlation of cPass with an ELISA detecting anti-RBD IgG, IgM, and IgA antibodies at a single timepoint and across intervals from onset of symptoms of SARS-CoV-2 infection.

**Results:** Compared to PRNT-50, cPass sensitivity ranged from 77% - 100% and specificity was 95% - 100%. Sensitivity was also high compared to the pseudotyped lentiviral neutralization assay (93% [95%CI 85-97]), but specificity was lower (58% [95%CI 48-67]). Highest agreement between cPass and ELISA was for anti-RBD IgG (*r*=0.823). Against the pseudotyped lentiviral neutralization assay, anti-RBD IgG sensitivity (99% [95%CI 94-100]) was very similar to that of cPass, but overall specificity was lower (37% [95%CI 28-47]). Against PRNT-50, results of cPass and anti-RBD IgG were nearly identical.

**Conclusions:** The added value of cPass compared to an IgG anti-RBD ELISA was modest.

## INTRODUCTION

Use cases for serological testing for prior exposure to *Severe acute respiratory syndrome-related coronavirus-2* (SARS-CoV-2) have been reviewed in detail (1, 2). Despite a rapid increase in the number and availability of serological assays detecting SARS-CoV-2 antibodies, critical knowledge gaps remain regarding the magnitude and kinetics of the correlation between results of these assays and the presence of neutralizing antibodies.

Only a subset of antibodies against a specific antigen can neutralize viral replication. Assays that measure neutralizing antibody levels, such as plaque reduction neutralization tests (PRNT) and microneutralization methods, provide essential data; these assays can help validate candidate diagnostic tests and define serological correlates of immunity. However, functional cell-based assays of SARS-CoV-2 neutralization can only be performed in a Biosafety Level 3 (BSL-3) laboratory, which is labour-intensive, costly, and severely limits testing throughput. Pseudotyped viruses have been developed that incorporate the Spike protein of SARS-CoV-2 and can be cultivated in BSL-2 conditions (3). Assays incorporating such pseudotyped viruses provide a functional assessment of the host neutralizing antibody responses as an alternative to using the wild-type virus (4-7). By contrast, surrogates of neutralization that bypass the need for viral culture would offer substantial advantages in terms of throughput, cost, and scalability. At least one direct ELISA assay detecting antibodies to the whole Spike protein has received regulatory approval in Europe for assessment of neutralizing antibodies (8). Further, several groups have proposed blocking assays, leveraging different signal detection methods to quantify the presence of host antibodies that can block the interaction of the SARS-CoV-2 Spike protein with human ACE-2 receptor (9-12).

On 6 Nov 2020, the FDA issued an emergency use authorization (EUA) for the cPass SARS-CoV-2 Neutralization Antibody Detection Kit (cPass; Genscript, Piscataway, NJ)(13), which is the first such surrogate neutralization assay to be commercially available. The cPass uses a blocking ELISA format with human ACE-2 receptor molecules coated on an ELISA plate (9, 14). Human sera pre-incubated with labelled epitopes of the receptor binding domain (RBD on S1 proteins) are then transferred to the plate. This blocking ELISA serves as a surrogate assay to inform on the capacity of human sera to block the interaction between the Spike fusion protein (through its RBD) and its cellular receptor ACE-2.

The objective of this study was to inform the use of the cPass and assess its added value compared to laboratory-developed anti-RBD ELISA assays by performing an evaluation using a variety of well characterised specimens. A number of reference panels were utilized to better understand the ability of the cPass assay to detect significant titres of neutralizing antibodies assessed by culture-based reference methods. We compared cPass to PRNT and to a pseudotyped virus neutralization assay. We also sought to describe the correlation of cPass with a laboratory-developed indirect ELISA detecting anti-RBD IgG, IgM, and IgA antibodies at a single timepoint and across different timeframes among specimens collected at a known interval from onset of symptoms of SARS-CoV-2 infection.

## METHODS

### Ethics

Research ethics board approval or exemption was obtained at all participating institutions.

### Source of specimens tested

We assembled several well-characterised SARS-CoV-2 serological specimen panels to assess the performance characteristics of the cPass culture-free neutralization antibody detection kit (Table 1). These panels included: a first panel from the Public Health Agency of Canada’s National Microbiology Laboratory comprising serological samples from COVID-19 patients, healthy individuals, as well as patients non-SARS-CoV-2 infections (NML panel 1; Supplemental Table 1); NML Panel 2 (the National SARS-CoV2 Serological Panel (NSSP)), comprising 60 serum or plasma specimens from persons with prior SARS-CoV-2 infection documented by nucleic acid amplification testing (NAAT) and 21 specimens from healthy blood donors collected in Canada prior to July 2019; the World Health Organization’s “First WHO International Reference Panel for anti-SARS-CoV-2 immunoglobulin” (NIBSC code 20/268) (15); and two separate curated panels from Héma-Québec and CR-CHUM. The later panels comprised convalescent plasma donors (confirmed SARS-CoV-2 infection and complete resolution of symptoms for at least 14 days) with either single timepoint or longitudinal follow-up. In addition to panels using neutralization assays as the reference standard, we assembled 136 specimens from healthy blood donors who tested negative for the presence of anti-SARS-CoV-2 antibodies by both a lab-developed anti-RBD IgG ELISA and a commercial assay detecting anti-nucleocapsid antibodies (Abbott Architect SARS-CoV-2 IgG Assay). These specimens, collected between 25 May and 9 July 2020, were acquired to help assess the ability of the cPass assay to detect specimens that test negative by other serological methods.

**TABLE 1.**
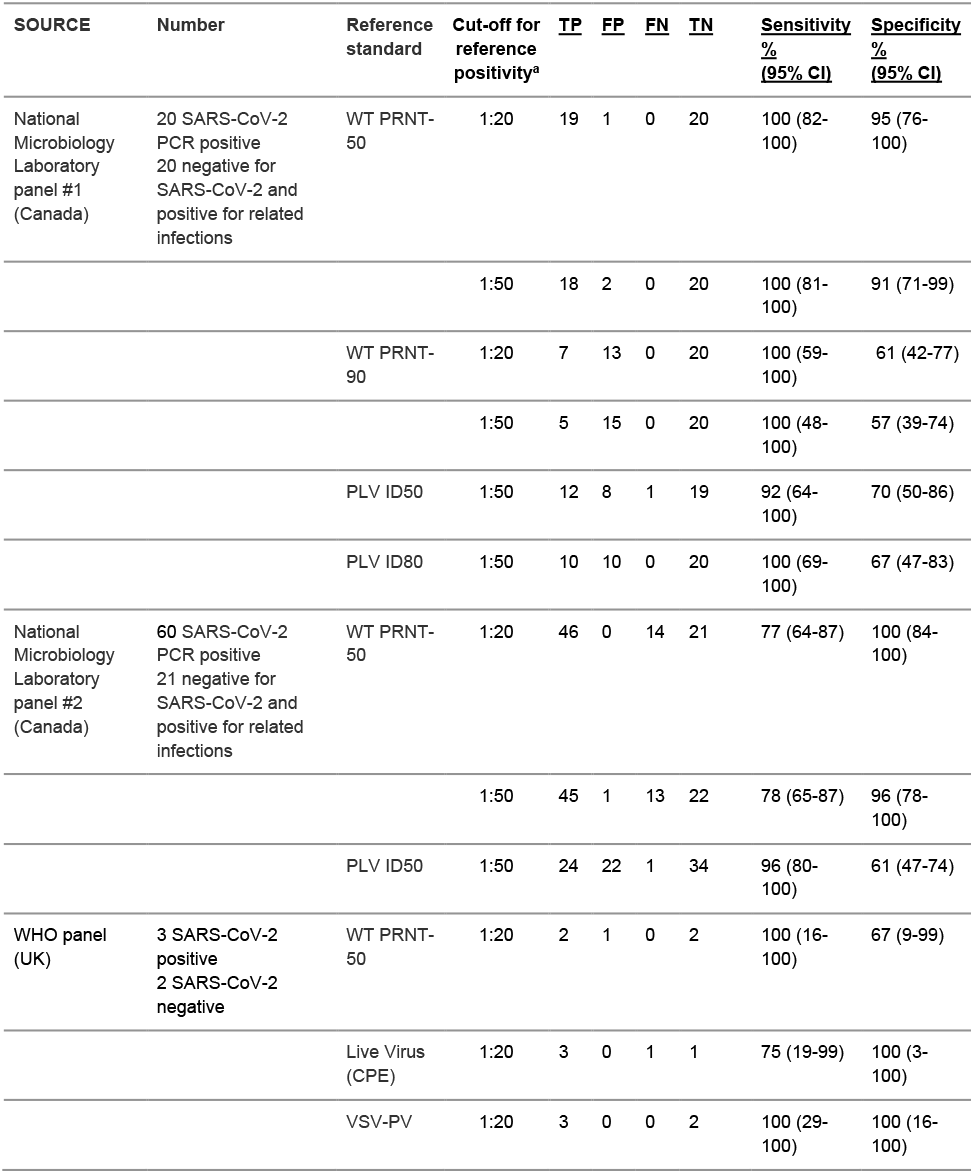
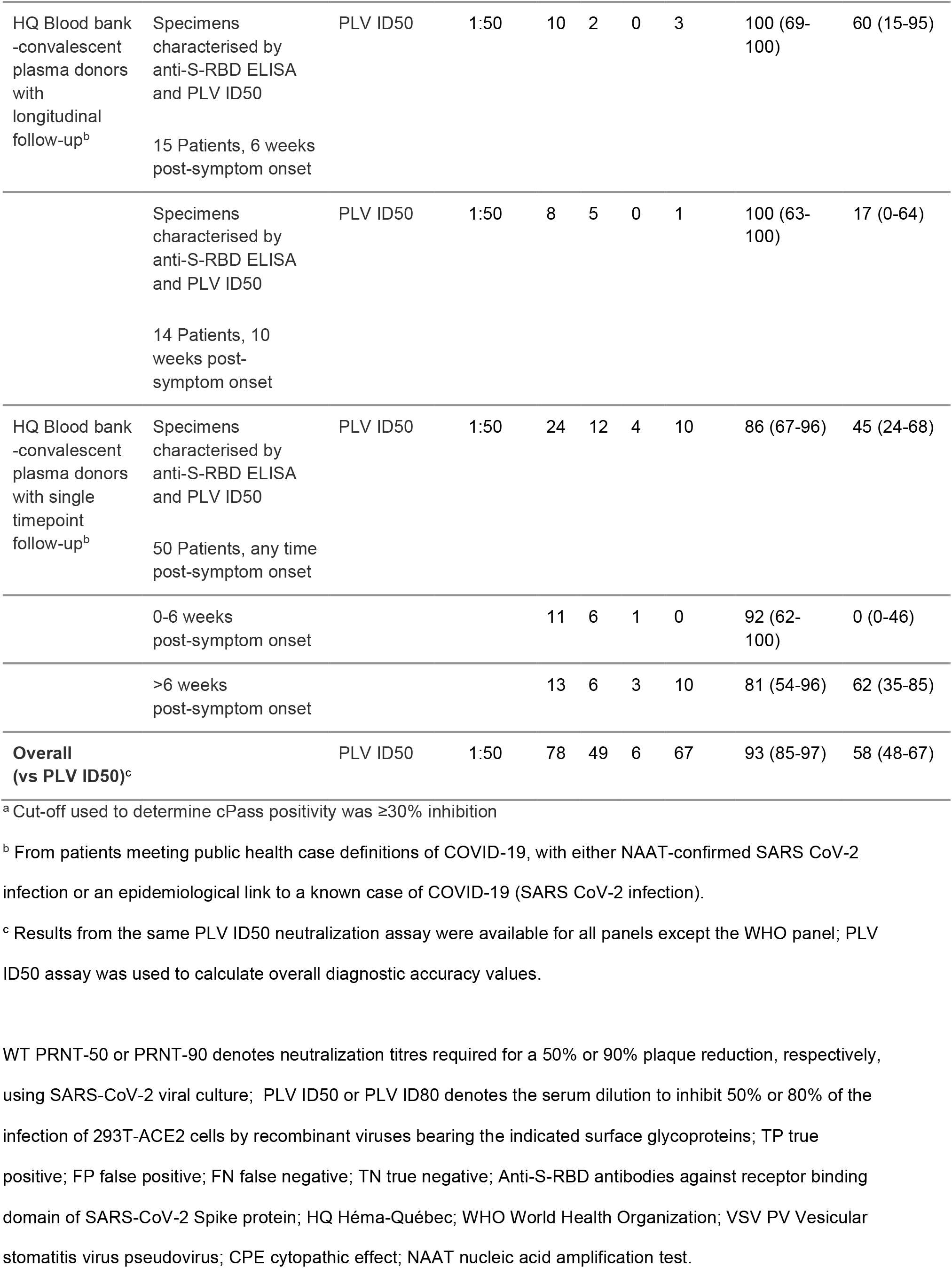
Diagnostic accuracy of the GenScript cPass surrogate viral neutralization assay to detect neutralizing antibodies among well-characterised specimen panels, according to reference standard used

### Culture-free neutralization antibody detection assay (cPass)

All specimens and controls were processed according to the manufacturer’s instructions (including a 10X dilution factor of the primary specimen) and were tested in triplicate. The percentage of inhibition calculation was based on the mean of OD for each triplicate. A cut-off of 30% inhibition of RBD-ACE2 binding was used to determine the presence of neutralizing antibodies, based on the manufacturer’s instructions for use.

### Detection of neutralizing antibodies by culture-based reference methods

Neutralizing antibodies were detected via either assessment of plaque reduction neutralization titres using wild-type SARS-CoV-2, or by determining the neutralization half-maximal inhibitory dilution (PLV ID50) or the neutralization 80% inhibitory dilution (PLV ID80) of pseudotyped lentiviral vector (16).

Assessment of plaque-reduction neutralization using wild-type SARS-CoV-2 was performed at the Public Health Agency of Canada’s National Reference Laboratory for Microbiology. Briefly, serological specimens were diluted 2-fold from 1:20 to 1:640 in DMEM supplemented with 2% FBS and challenged with 50 plaque forming units (PFU) of SARS-CoV-2 (hCoV-19/Canada/ON_ON-VIDO-01-2/2020, EPI_-ISL_425177), which were titrated by plaque assay (17). After 1 hour of incubation at 37°C and 5% CO2, the sera-virus mixtures were added to 12-well plates containing Vero E6 cells at 90% to 100% confluence and incubated at 37°C and 5% CO2 for 1 hour. After adsorption, a liquid overlay comprising 1.5% carboxymethylcellulose diluted in MEM supplemented with 4% FBS, L-glutamine, non-essential amino acids, and sodium bicarbonate was added to each well and plates were incubated at 37°C and 5% CO2 for 72 hours. The liquid overlay was removed, and cells were fixed with 10% neutral-buffered formalin for 1 hour at room temperature. The monolayers were stained with 0.5% crystal violet for 10 minutes and washed with 20% ethanol. Plaques were enumerated and compared to controls. The highest serum dilution resulting in 50% and 90% reduction in plaques compared with controls were defined as the PRNT-50 and PRNT-90 endpoint titres, respectively. PRNT-50 titres and PRNT-90 titres ≥1:20 were considered positive for SARS-CoV-2 neutralizing antibodies.

Pseudoviral neutralization testing was performed as previously described (16). Briefly, target cells were infected with single-round luciferase-expressing lentiviral particles. HEK 293T cells were transfected by the calcium phosphate method with the lentiviral vector pNL4.3 R-E-Luc (NIH AIDS Reagent Program) and a plasmid encoding for SARS-CoV-2 Spike at a ratio of 5:4. Two days post-transfection, cell supernatants were harvested and stored at –80°C until use. 293T-ACE2 target cells were seeded at a density of 1 × 10^4^ cells/well in 96-well luminometer-compatible tissue culture plates (Perkin Elmer) 24h before infection. Recombinant viruses in a final volume of 100 µL were incubated with the indicated sera dilutions (1/50; 1/250; 1/1250; 1/6250; 1/31250) for 1h at 37°C and were then added to the target cells followed by incubation for 48h at 37°C; cells were lysed by the addition of 30 µL of passive lysis buffer (Promega) followed by one freeze-thaw cycle. An LB942 TriStar luminometer (Berthold Technologies) was used to measure the luciferase activity of each well after the addition of 100 µL of luciferin buffer (15mM MgSO_4_, 15mM KPO_4_ [pH 7.8], 1mM ATP, and 1mM dithiothreitol) and 50 µL of 1mM d-luciferin potassium salt (ThermoFisher Scientific). The neutralization half-maximal inhibitory dilution (ID50) or the neutralization 80% inhibitory dilution (ID80) represents the sera dilution to inhibit 50% or 80% of the infection of 293T-ACE2 cells by recombinant viruses bearing the indicated surface glycoproteins.

### Indirect anti-RBD ELISA assays

Specimens were analysed with a laboratory-developed indirect ELISA detecting anti-RBD IgG, IgM, and IgA as previously described (16, 18).

### Statistical analysis

The diagnostic accuracy of the cPass surrogate viral neutralization assay was estimated compared to different reference standards (WT PRNT-50; WT PRNT-90; PLV ID50; PLV ID80, Live Virus (CPE), and VSV-PV). Sensitivities and specificities are presented with 95% confidence intervals (95% CI). The effect of varying the cut-off value (i.e., % inhibition of RBD-ACE2 binding) for cPass positivity on the diagnostic accuracy of the cPass against a PLV PRNT-50 reference standard was investigated using a receiver operating characteristic (ROC) curve. The association between cPass % inhibition and results obtained using laboratory-developed ELISA detecting anti-S-RBD IgG, IgM, and IgA are presented in scatterplots with the strength of these associations informed by Pearson correlation. Lastly, among specimens from individuals with a known interval from onset of SARS-CoV-2 infection symptoms and repeated testing over time, spaghetti plots were created to investigate any change in signal over time for the cPass and direct anti-S-RBD ELISA with statistical significance assessed using the Wilcoxon matched-pairs signed-rank test (p<0.05 denoted by *). Statistical analyses were performed using R version 3.5.2 (R Core Team, Vienna, Austria).

## RESULTS

### Diagnostic accuracy for the detection of anti-SARS-CoV-2 neutralizing antibodies, and the impact of using different reference standards

Table 1 shows the estimated diagnostic accuracy of the GenScript cPass neutralization antibody detection assay among well characterised specimen panels, according to different reference standards. Among various reference standards, results from the same PLV ID50 assay were available for all panels except the WHO panel, and this was used to estimate aggregate diagnostic accuracy values across several panels.

Overall, cPass had sensitivity ranging 77% - 100% and specificity of 95% - 100% compared to the reference standard of a 50% plaque reduction neutralization using SARS-CoV-2 viral culture (WT PRNT-50) (Table 1). Changing the WT PRNT-50 cut-off titre from 1:20 to 1:50 had minimal impact on specimen categorization. Sensitivity remained very high compared to the reference standard of a neutralization half-maximal inhibitory dilution using a validated pseudotyped lentiviral vector neutralization assay (PLV ID50) with a cut-off titre of 1:50, but specificity was lower than that compared to WT PRNT-50, ranging from 17-70% (Table 1).

The effect of cut-off values on the diagnostic accuracy of the GenScript cPass assay is shown in Figure 1. A receiver operating characteristic (ROC) curve using the reference standard of PLV ID50 yielded an area under the ROC curve of 0.858.

**Figure 1.**
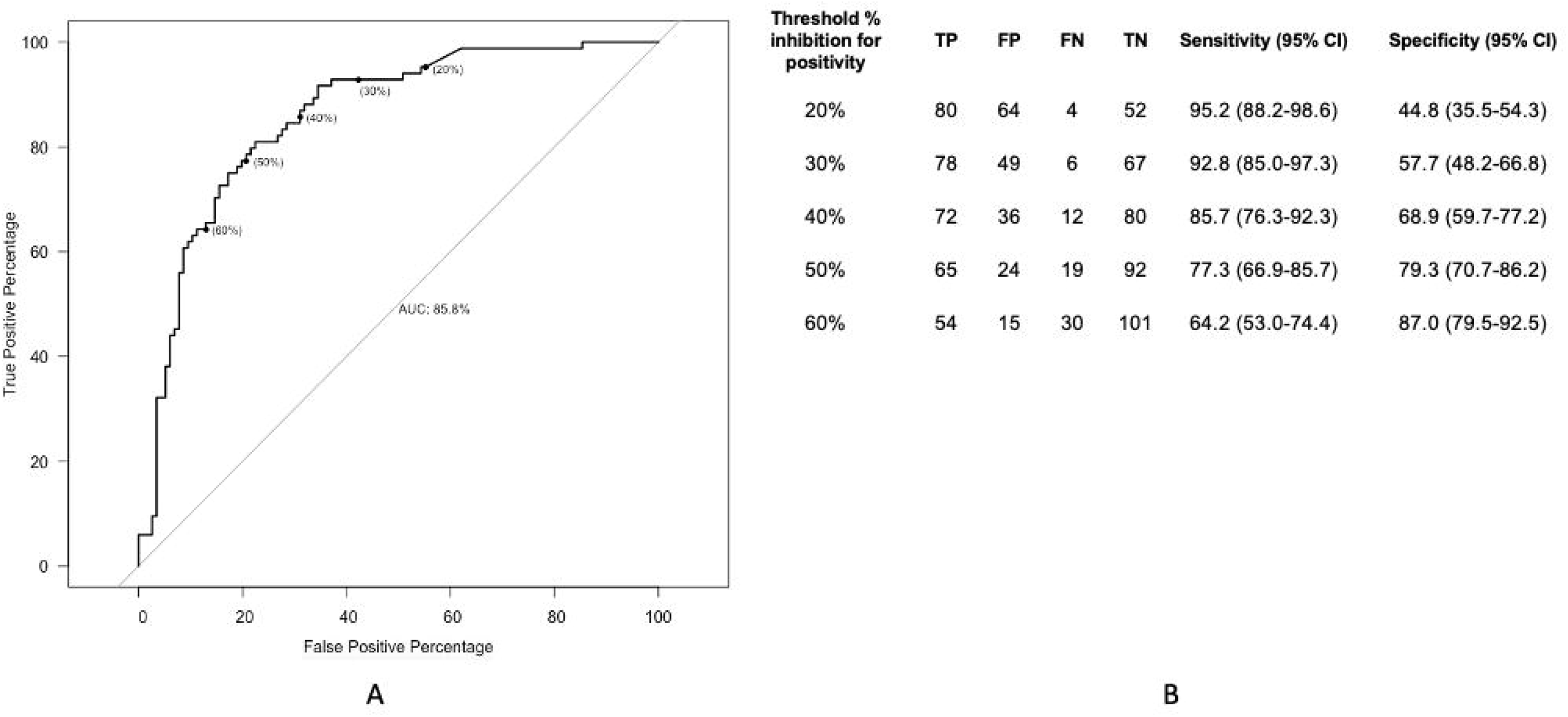
Effect of cut-off values on the diagnostic accuracy of the Genscript cPass SARS-CoV-2 neutralization antibody detection kit. Panel (A) shows the receiver operating characteristic (ROC) curve, with different cPass cut-offs. Panel (B) details results and estimates of sensitivity and specificity for different %inhibition of RBD-ACE2 binding cut-offs for cPass positivity. The reference standard used is PLV PRNT 50 at a titre of ≥1:50. The specimens from the WHO panel (n=5) are not included in the above figure as PLV PRNT 50 was not performed, thereby resulting in a total N of 200. AUC denotes Area Under the ROC Curve; TP true positive; FP false positive; FN false negative; TN true negative.

### Effect of serial dilution on the accuracy for detecting sera with positive PRNT-90 titres

Against the most stringent reference standard of 90% plaque reduction neutralization using SARS-CoV-2 viral culture (WT PRNT-90), estimated specificity was reduced compared to WT PRNT-50. Specificity remained similar whether a cut-off WT PRNT-90 titre for positivity of 1:20 or 1:50 was used [61% (95%CI 42-77) and 57% (95%CI 39-74), respectively] (Table 1). We performed serial dilution of the 16 primary specimens from the National Microbiology Laboratory Panel with WT PRNT-50 titres ≥1:20 to determine whether we could establish a dilution that increased specificity for detecting those with WT PRNT-90 titres ≥1:20 without sacrificing sensitivity (Figure 2). A 50-fold dilution of specimens with positive WT PRNT-50 titres increased specificity for those with positive WT PRNT-90 titres from 11% (95%CI 0-48) to 100% (95%CI 66-100), with one missed PRNT-90 positive specimen.

**Figure 2.**
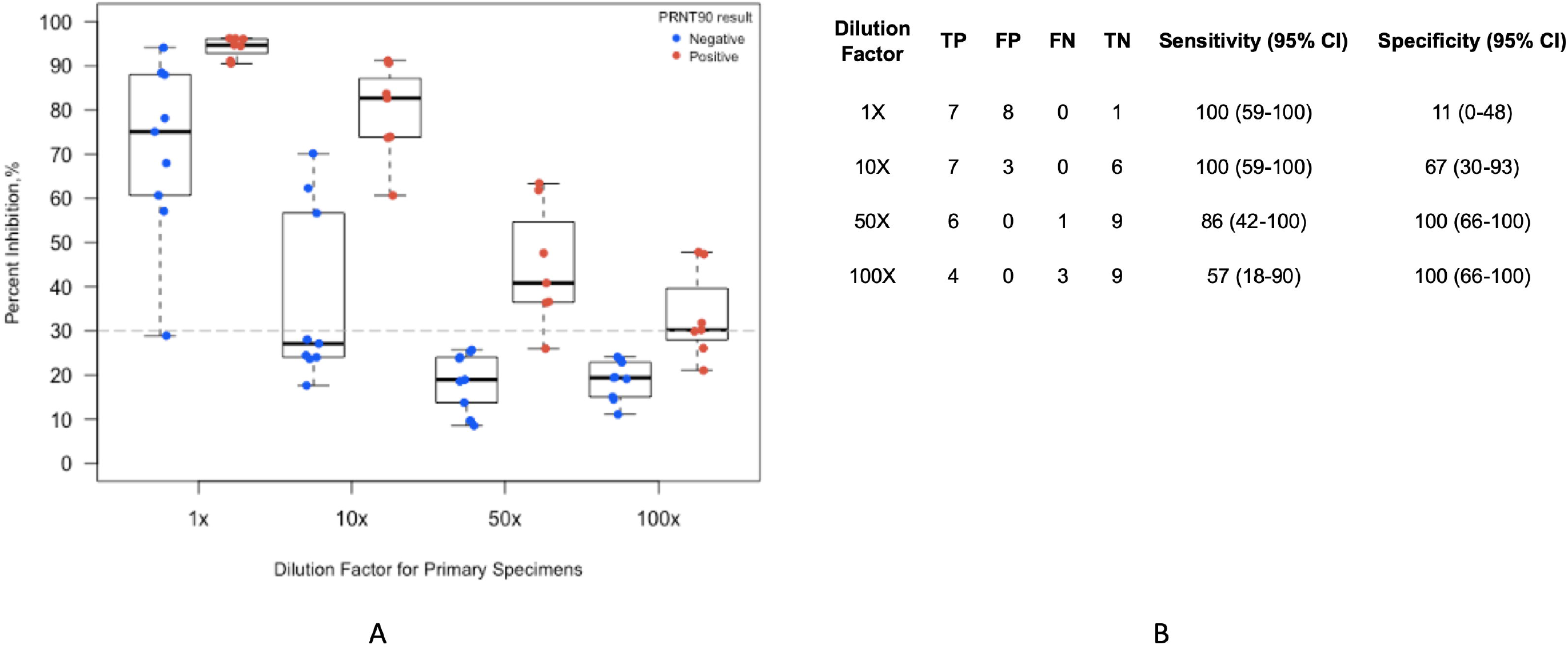
Effect of serial dilution on the accuracy for detecting sera with positive PRNT90 titres. Serial dilution of the 16 primary specimens with WT PRNT 50 titres ≥1:20 was performed to establish a dilution that increased specificity for detecting those with WT PRNT 90 titres ≥1:20. Panel (A) shows individual data points according to dilution and WT PRNT 90 status (positive ≥1:20). Box plots depict the median and interquartile range. Panel (B) details results and estimates of sensitivity and specificity for serial dilution factor. All dilution factors are additional to the 10X dilution required in the manufacturer’s instructions. WT PRNT 90 denotes neutralization titres required for a 90% plaque reduction using SARS-CoV-2 viral culture; TP true positive; FP false positive; FN false negative; TN true negative.

### Agreement of the GenScript cPass assay with laboratory-developed ELISA detecting anti-RBD IgG, IgM, and IgA

Results obtained with cPass were compared to those obtained using laboratory-developed ELISA detecting anti-RBD IgG, IgM, and IgA to assess whether the cPass yields complementary information (Figure 3). Highest agreement between cPass percent inhibition of RBD-ACE2 binding and ELISA readout was seen for anti-RBD IgG (Pearson correlation coefficitient *r*=0.823), compared to that observed with anti-RBD IgM and IgA (*r*=0.505 and 0.489, respectively). The diagnostic accuracy of categorical anti-RBD IgG results for the detection of SARS-CoV-2 neutralizing antibodies was very similar to that observed with the cPass for most panels and reference standards (Tables 1 and 2). Compared to PLV ID50, cPass overall sensitivity was 93% [95% CI 85-97] and specificity 58% [95% CI 48-67], whereas anti-RBD IgG overall sensitivity was 99% [95% CI 94-100] and specificity 37% [95% CI 28-47].

**TABLE 2.**
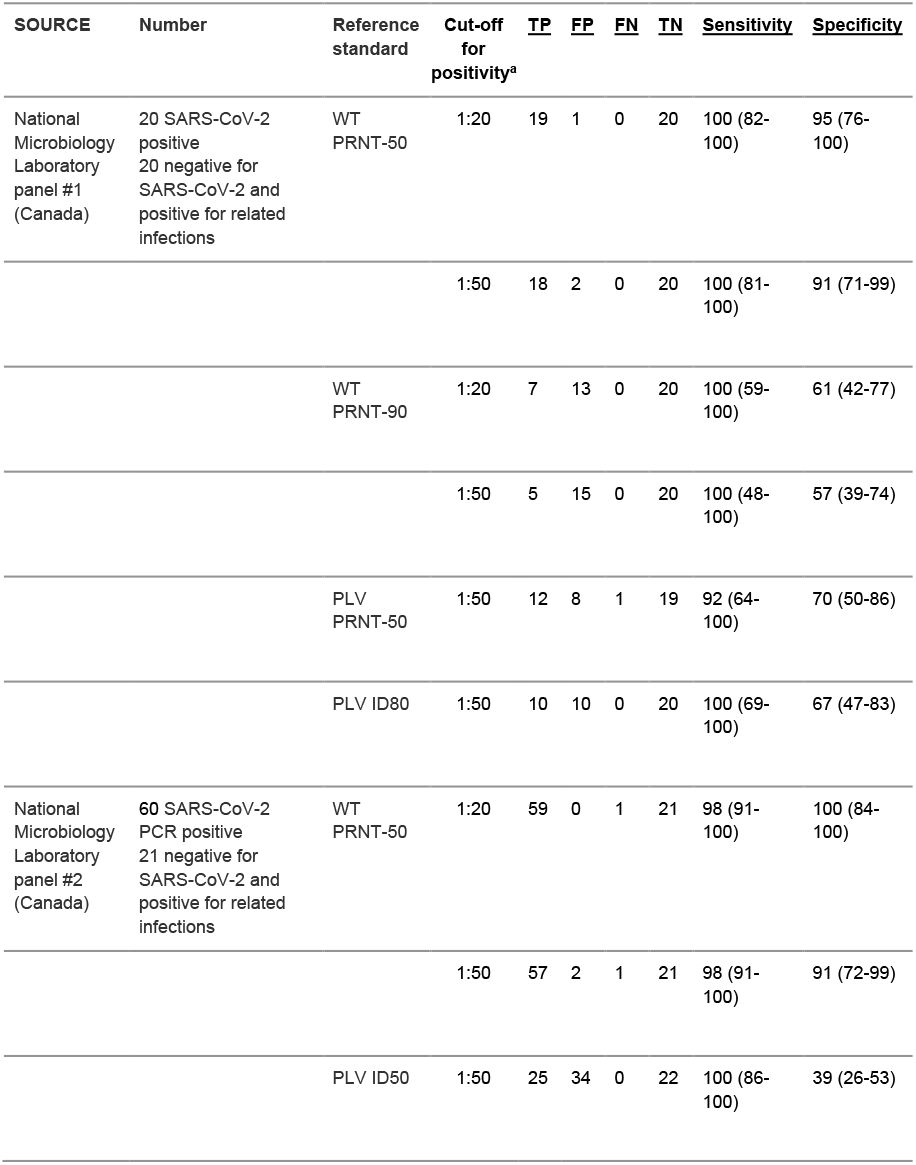
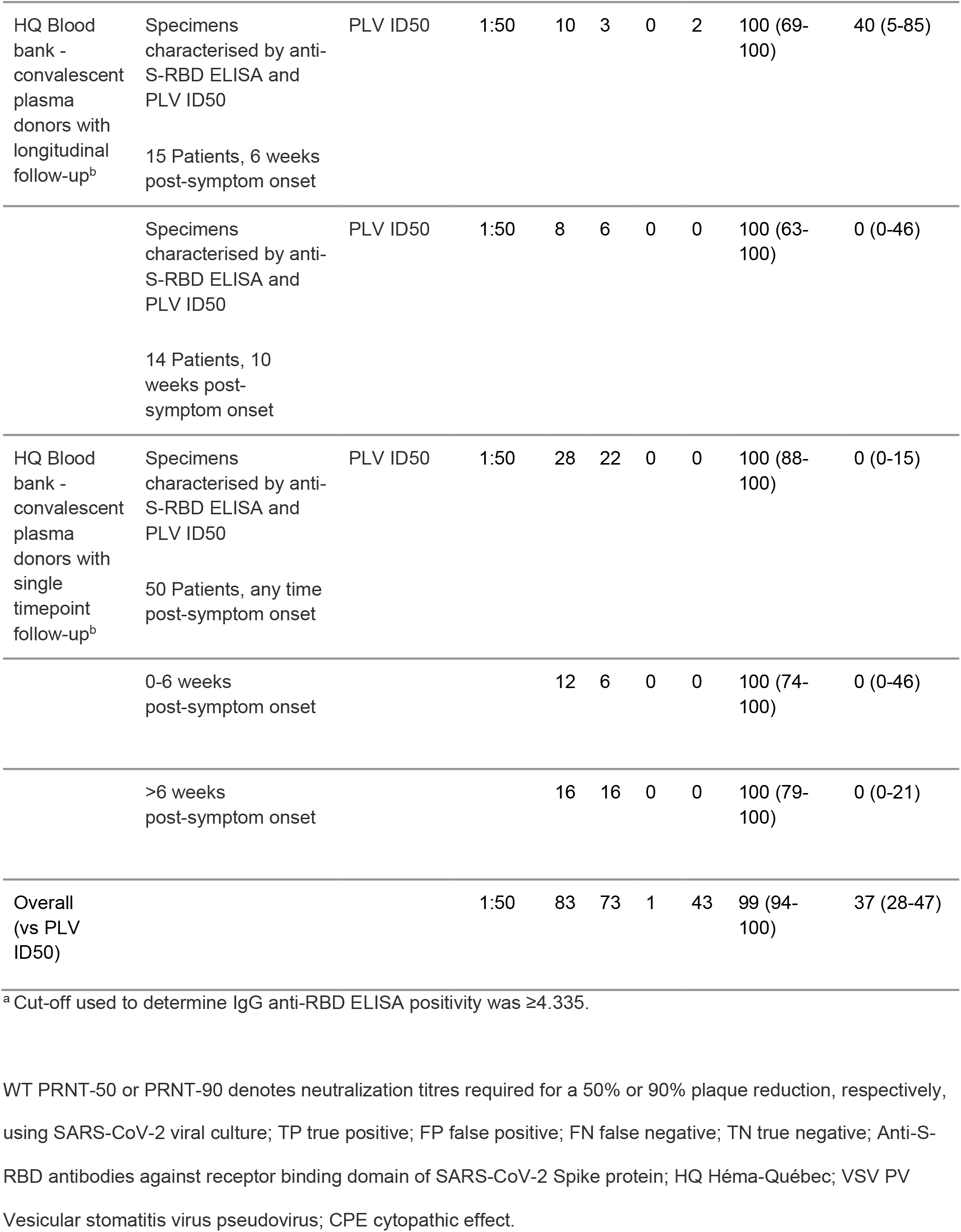
Diagnostic accuracy of a laboratory-developed IgG anti-RBD ELISA to detect neutralizing antibodies

**Figure 3.**
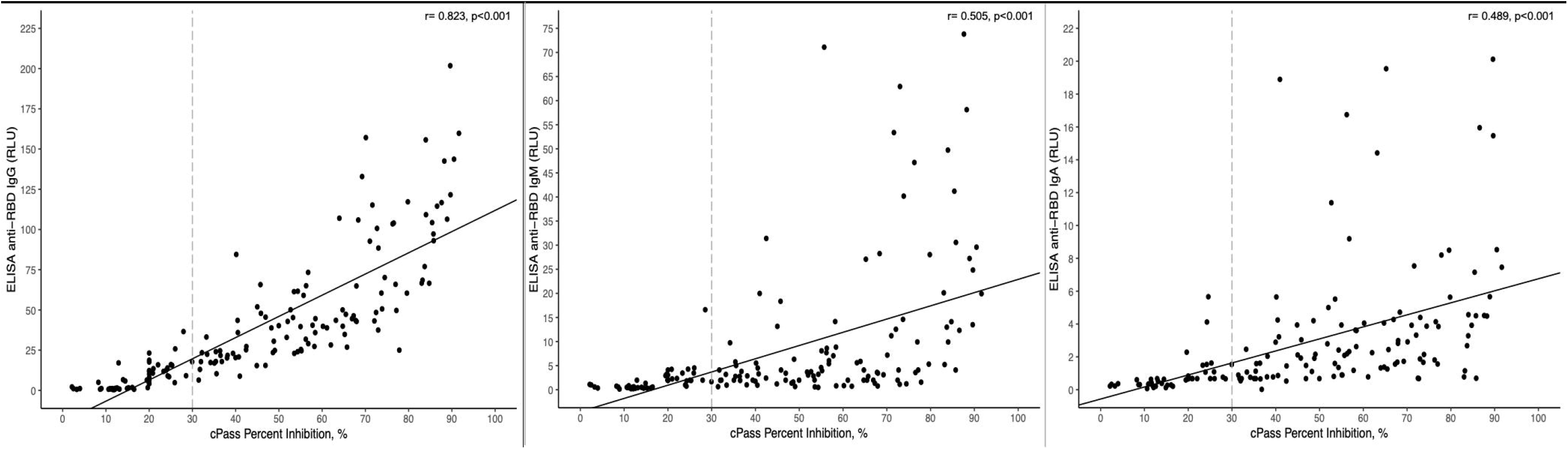
Correlation of the Genscript cPass assay with anti-S-RBD ELISA. Correlation of the Genscript cPass assay with the anti-S-RBD ELISA normalized relative luciferase units (RLU) for each plasma tested at a dilution (1:500) is presented. Scatterplots and Pearson correlation coefficient for results obtained with cPass compared to those obtained using laboratory-developed ELISA detecting anti-RBD IgG, IgM, and IgA (Panels A, B, C, respectively). The vertical dashed line depicts the manufacturer’s recommended cut-off for cPass positivity. Specimens from the NML panel 2 and Héma-Québec convalescent plasma donors panel are included in the above figure. Specimens from the NML panel #1 were excluded as anti-S-RBD ELISA for IgM and IgA were not performed.

However, when NML panel 2 was considered in isolation, categorical results for the detection of SARS-CoV-2 neutralizing antibodies differed substantially between cPass and anti-RBD IgG in terms of sensitivity compared to WT PRNT-50 (cPass 77% [95% CI 64-87], anti-RBD IgG 98% [95% CI 91-100]) and specificity compared to PLV ID50 (cPass 61% [95% CI 47-74], anti-RBD IgG 39% [95% CI 26-53]). If a cut-off of 20% RBD-ACE2 binding inhibition were used instead of the 30% cut-off recommended by the manufacturer, cPass sensitivity against WT PRNT-50 would rise to 92% [95% CI 82-97] with a lower estimated specificity of 46% [95% CI 33-60].

Among paired specimens from the same individual collected at a known interval from SARS-CoV-2 diagnosis, aggregate results of both cPass and direct anti-RBD IgG ELISA did not change between 6 weeks and 10 weeks after diagnosis (p=1.00 and 0.104, respectively, by the Wilcoxon signed rank test) (Figure 4). In contrast, ELISA readouts decreased significantly over the same timeframe for direct anti-RBD IgM (p=0.0058) and IgA (p=0.0012).

**Figure 4.**
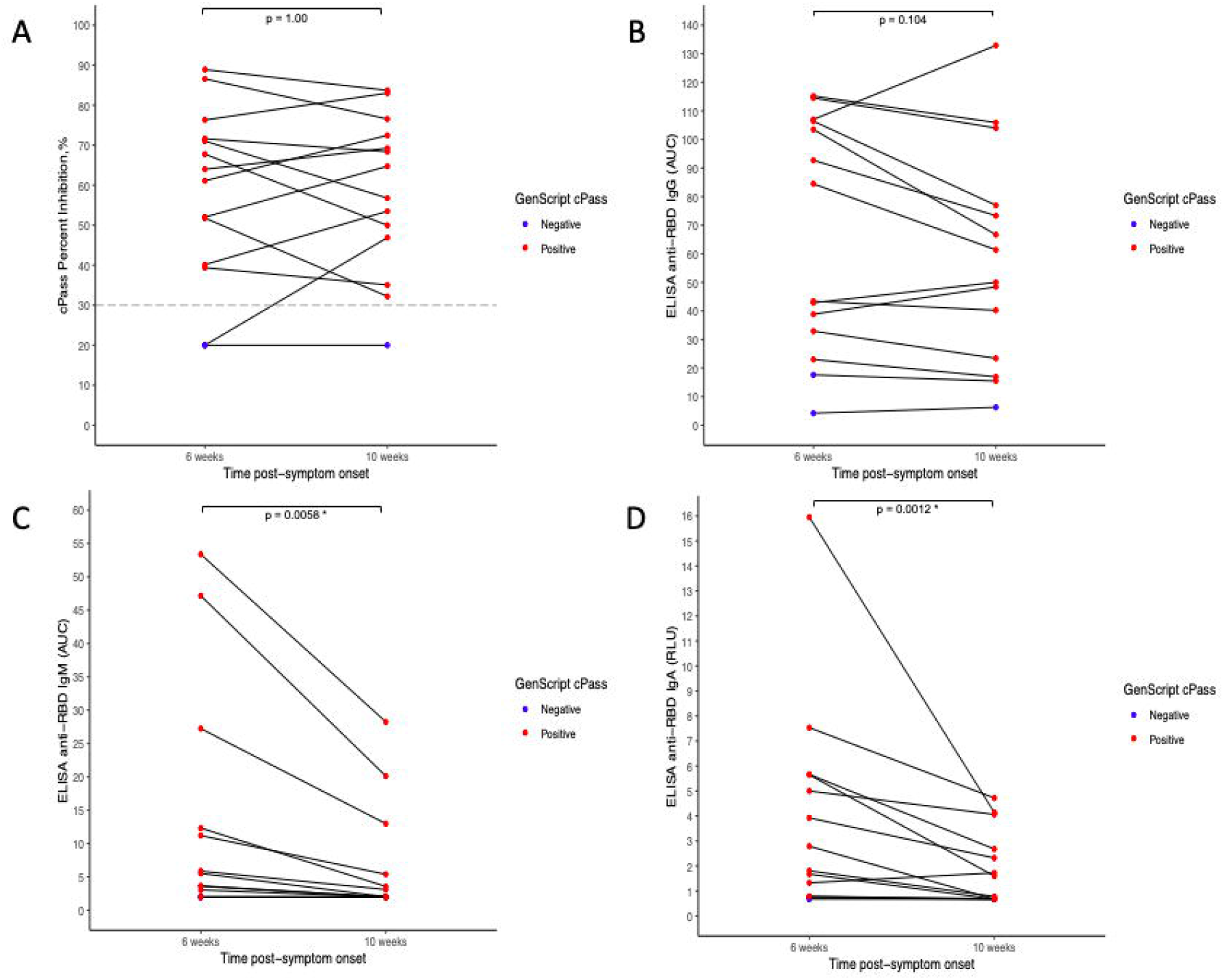
Change of signal over time for Genscript cPass and anti-RBD ELISA. Spaghetti plot of results obtained with cPass (panel A) and the plots shown in panels B, C, D represent (B and C) the areas under the curve (AUC) calculated from relative luciferase units (RLU) obtained with serial plasma dilutions or (D) the normalized RLU for one plasma dilution (1:500) for laboratory-developed ELISA detecting anti-RBD IgG, IgM, and IgA (panels B, C, D, respectively) among specimens collected at a known interval from SARS-CoV-2 diagnosis. Horizontal lines indicate paired specimens form the same individual. P values are calculated via the Wilcoxon signed rank test, and values <0.05 are designated with an Asterix. In all panels, red dots denote specimens with positive cPass results, and blue dots specimens with negative cPass results.

### Negative agreement between cPass and other serological assays

Among 136 specimens from healthy blood donors who tested negative for the presence of anti-SARS-CoV-2 antibodies by both a lab-developed anti-RBD IgG ELISA and the Abbott Architect SARS-CoV-2 IgG Assay (anti-N protein), cPass yielded negative results for 134 specimens (negative agreement 98.5% [95% CI 94.8 – 99.8]).

## DISCUSSION

Rapid and high throughput surrogates for PRNT or pseudovirus neutralization assays that bypass the need for cell culture are awaited with the belief that they will offer additional information to that from standard direct immunoassays, such as a higher specificity for neutralizing antibodies. The cPass SARS-CoV-2 Neutralization Antibody Detection Kit (cPass) is the first such assay to be commercially available and to receive FDA EUA in the U.S. An evaluation of a cPass prototype, using a cut-off value of 20% inhibition, found that it could provide a high-throughput screening tool for confirmatory PRNT testing (19). The results of the current evaluation support the ability for cPass to detect neutralizing antibodies to SARS-CoV-2, although specificity varied considerably depending on the reference assay used. Our data also extend these findings by showing that cPass performed similarly to a non-blocking anti-RBD ELISA among varied well characterised specimen panels.

Among 205 specimens evaluated by a SARS-CoV-2 reference neutralization assay in the current work - either WT PRNT-50 or PLV ID50 - the overall estimated sensitivity of cPass for detection of anti-SARS-CoV-2 neutralizing antibodies was high, regardless of the reference standard technique or reference standard cut-off titre for positivity. The lower sensitivity of cPass compared to WT PRNT-50 observed for specimens in NML panel 2 (Table 1) appears related to the choice of 30% RBD-ACE2 binding inhibition cut-off recommended by the manufacturer, which may result in false negative results for specimens with low titres of neutralizing antibodies. Among all specimens evaluated, however, reducing the inhibition cut-off to 20% would have a minimal impact on overall sensitivity and yield substantial reduction in overall specificity compared to PLV-50 (Figure 1). Our results do not suggest that the cPass assay, targeting only RBD-ACE2 blockade, would miss a substantial proportion of patients with neutralizing antibodies that target non-RBD epitopes (20-22). This may be because neutralizing antibodies to non-RBD epitopes usually occur concomitantly with anti-RDB neutralizing antibodies, instead of in isolation.

By contrast, estimates of the specificity of cPass for the detection of anti-SARS-CoV-2 neutralizing antibodies were contingent of the reference standard used (Table 1). There was near-perfect negative agreement with WT PRNT-50 using a cut-off titre of either 1:20 or 1:50. However, negative agreement was much lower when cPass was compared to either PLV ID50 or WT PRNT-90. Our data raise the unresolved questions of which reference technique (i.e., wild-type or pseudotyped live viral culture), level of stringency (e.g., 50% inhibition of infection vs 80%, 90%, etc), and cut-off titre (e.g., 1:20 vs 1:50) best represent serocorrelates of protection to SARS-CoV-2, or other relevant applications. Moreover, protocols can vary widely for the same technique across different laboratories, requiring caution in the interpretation of these and other data (23). In the current manuscript, PLV ID50 with a cut-off titre of 1:50 was used as the overall comparator because it was the technique applied to all available specimen panels except the 5-member panel from WHO. Our results must be interpreted in context with this potential source of bias. However, we note that this technique has been employed by other groups and thus offers a high degree of generalizability with other results (24, 25).

The cPass assay detected all specimens with positive WT PRNT-90 titres, with a significant proportion of false positives (Figure 2). A 50-fold dilution of the 16 primary specimens with WT PRNT-50 titres ≥1:20 increased specificity for detecting those with WT PRNT-90 titres ≥1:20 from 11% (95% CI 0-48) to 100% (95% CI 66-100). This may represent a useful approach for using the cPass assay to identify blood specimens with positive WT PRNT-90 titres, which has been proposed as a desirable characteristic for sera used in convalescent plasma trials by some regulatory agencies.

Finally, results of the cPass assay are best correlated with those of a laboratory-developed indirect anti-RBD ELISA detecting IgG, both at a single timepoint (Table 2, Figure 3) and across time among paired specimens form the same individual collected at a known interval from symptoms onset (Figure 4). However, a slightly higher specificity for the detection of anti-SARS-CoV-2 neutralizing antibodies was observed for cPass compared to anti-RBD IgG ELISA across most panels (Tables 1 and 2). The fact that results of cPass and anti-RBD IgG remained stable between 6 and 10 weeks post-symptom onset, while ELISA readouts decreased significantly over the same timeframe for anti-RBD IgM and IgA is potentially concerning given recent work suggesting a major role of IgM and IgA in the neutralizing activity of convalescent plasma against SARS-CoV-2 (18, 26-28). The observed trend toward lower specificity of cPass at later timepoints among convalescent plasma donors with longitudinal follow-up (i.e. [60% (95% CI 15-95)] at 6 weeks vs [17% (95% CI 0-64)] at 10 weeks) may thus be related to loss of neutralizing IgM (Table 1). Taken together, these results suggest that a positive cPass result in the context of a remote infection may not accurately predict the presence of neutralizing antibodies. In addition, specificity of the cPass may be affected by the possibility that part of the inhibition of binding in the cPass assay could be due to steric hindrance by the abundant anti-Spike antibodies of the IgG isotype rather than by true neutralization (as occurs in vivo).

## CONCLUSIONS

The results of the current evaluation demonstrate the ability of cPass to detect blood specimens with anti-SARS-CoV-2 neutralizing antibodies. However, the added value of cPass compared to an IgG anti-RBD ELISA was modest.

## Data Availability

After peer-reviewed publication, data available on request in the setting of a suitable research protocol.

## ACKNOWLEDGEMENTS

We thank the convalescent plasma donors who participated in this study; the Héma-Québec team involved in convalescent donor recruitment and plasma collection; the staff members of the CRCHUM BSL3 Platform for technical assistance; Stefan Pöhlmann (Georg-August University, Germany) for the plasmid coding for SARS-CoV-2 S.

