## Supplemental Table 1 for "Evaluation of a Commercial Culture-free Neutralization Antibody Detection Kit for Severe Acute Respiratory Syndrome-Related Coronavirus-2 and Comparison with an Anti-RBD ELISA Assay"

**Supplementary Table 1.** Composition of SARS-CoV-2 negative serum samples in National Microbiology Laboratory panel

| Infections among the SARS-COV-2 negative samples provided by NML | Number of negative NML samples n, (%) |
|---|---|
| Hepatitis | 5 (25%) |
| HIV | 3 (15%) |
| Rabies | 5 (25%) |
| SARS-1 | 2 (10%) |
| Syphilis | 5 (25%) |

NML, National Microbiology Laboratory
